# Prevalence and Antimicrobial Resistance of *Salmonella* spp., *Shigella* spp., and *Listeria monocytogenes* in Poultry Feeds and Ready-to-Eat Foods: A Farm-to-Fork Study in Conflict-Affected Maiduguri, Nigeria

**DOI:** 10.64898/2026.03.20.26348892

**Authors:** H.B. Ali, Barka Abbas Maladi, Farida Abdullahi Alhassan, Jummai James Bwalla, Isyaka M. Tom, J.M Ajagbe, M Usman, Abubakar Sadiq Baba, Yahaya Usman

**Affiliations:** Department of Medical Laboratory Science, Faculty of Allied Health Sciences University of Maiduguri Borno State, Nigeria; Department of Medical Laboratory Science, Federal University of Applied Sciences Kachia, Kaduna State; Department of Medical Microbiology Faculty of Basic Clinical Sciences College of Medical Sciences University of Maiduguri; Department of Medical Laboratory Sciences, College of Medical Sciences, Ahmadu Bello University, Zaria, Kaduna State, Nigeria

**Keywords:** *Salmonella* spp., *Shigella* spp., *Listeria monocytogenes*, farm-to-fork transmission, antimicrobial resistance, poultry feeds, ready-to-eat foods, Maiduguri, conflict-affected Nigeria, food safety

## Abstract

**Background:** Informal food supply chains in regions affected by conflict present significant risks associated with the transmission of foodborne pathogens and the development of antimicrobial resistance (AMR). The objective of this research is to investigate the prevalence and AMR patterns of *Salmonella* spp., *Shigella* spp., and *Listeria monocytogenes* in poultry feeds and ready-to-eat (RTE) foods available for sale in Maiduguri, located in northeastern Nigeria.

**Methods:** A cross-sectional survey was conducted involving the collection of 120 poultry feed samples and approximately 251 samples of RTE foods, with 120 samples specifically designated for the analysis of Listeria. Pathogen isolation was performed using standardized microbiological protocols (ISO/FDA-BAM), and the assessment of AMR was conducted utilizing the disk diffusion method in accordance with Clinical and Laboratory Standards Institute (CLSI) guidelines for *Shigella* (n=5), *Salmonella* (n=15), and Listeria (n=3) isolates. Prevalence rates were calculated with 95% confidence intervals derived from the Wilson score method.

**Results:** The study revealed a prevalence of *Salmonella* spp. at 10.0% (95% CI 5.8–16.8) in poultry feeds, with a maximum occurrence of 25.0% identified in the Monday Market, contrasting with a prevalence of 2.3% (0.8–6.7) reported in RTE foods, particularly in Bulumkutu fruits and meats from the Custom Market. *Shigella* spp. was identified in 3.33% (1.3–8.1) of feed samples and in 0.76% (0.1–4.2) of RTE foods, specifically within pineapple. *L. monocytogenes* was detected in 2.5% (0.8–7.1) of RTE foods, exclusively sourced from Baga Road market, including meat and sala sour milk. The isolates of *Salmonella* obtained from feeds demonstrated high levels of multidrug resistance (100% to tetracycline and 83.33% to fluoroquinolones), while *Salmonella* from RTE foods maintained resistance to tetracycline but exhibited a loss of resistance to fluoroquinolones. Notably, *L. monocytogenes* exhibited 100% resistance to both fluoroquinolones and cephalosporins while remaining sensitive to tetracycline. The presence of *coliforms* indicated inadequate hygiene conditions in the feeds, with 60.0% of samples showing contamination.

**Conclusion:** This initial study, which encompasses multiple pathogens within the conflict-affected region of Maiduguri, reveals significant upstream contamination of *Salmonella* in poultry feeds. It delineates the risks associated with ready-to-eat (RTE) foods and highlights the exclusive presence of *L. monocytogenes* in RTE products. Furthermore, the study indicates divergence in antimicrobial resistance (AMR) based on the food matrix. The findings underscore the urgent need for enhanced One Health surveillance, the implementation of improved hygiene practices in feed production, and the regulation of antimicrobial usage in informal poultry systems. Such measures are essential to effectively mitigate foodborne threats and the emergence of antimicrobial resistance in similarly underserved regions.

## 1.0 Introduction

Foodborne diseases continue to pose a significant public health challenge on a global level, with recent estimates indicating that unsafe food is accountable for approximately 600 million illnesses and 420,000 fatalities each year. Low- and middle-income countries bear the most substantial burden of these health issues [1]. In sub-Saharan Africa, the risks are intensified by informal food systems, inadequate infrastructure, and ongoing conflicts, particularly within urban centers that rely heavily on street vendors and ready-to-eat (RTE) food offerings [2]. Maiduguri, the capital of Borno State in northeastern Nigeria, serves as a compelling example of such a high-risk environment, where persistent security challenges have disrupted supply chains, increased dependence on informal markets, and limited access to effective food safety surveillance [3].

*Salmonella* spp., *Shigella* spp., and *Listeria monocytogenes* represent some of the most significant bacterial pathogens associated with foodborne illnesses. *Salmonella* spp., a Gram-negative organism belonging to the Enterobacteriaceae family, encompasses over 2,500 serovars and is a primary cause of zoonotic gastroenteritis, frequently linked to contaminated poultry products and feeds [4]. *Shigella* spp., also a member of the Enterobacteriaceae family, is responsible for shigellosis, an acute diarrheal disease that is transmitted via the fecal-oral route through contaminated food, water, or direct contact [5]. *Listeria monocytogenes*, classified as a Gram-positive facultative intracellular pathogen, poses considerable risks in the context of RTE foods due to its capacity to survive refrigeration, which may result in severe invasive listeriosis, particularly among susceptible populations [6]. The emergence of antimicrobial resistance (AMR) in these pathogens represents an escalating threat, as multidrug-resistant strains complicate treatment and contribute to increased morbidity and mortality rates in resource-limited settings [7].

The Global Antimicrobial Resistance and Use Surveillance System (GLASS), established by the World Health Organization in 2015, offers a standardized framework for monitoring trends in antimicrobial resistance (AMR) among priority bacterial pathogens associated with human infections. The system’s scope is progressively expanding to encompass antimicrobial consumption and One Health approaches pertinent to foodborne transmission [22]. Despite the significant burden posed by foodborne diseases in Nigeria, there is a notable deficiency of studies examining the transmission of these pathogens from production to consumption in poultry feeds and RTE foods, particularly in conflict-affected northeastern regions where informal markets dominate and regulatory oversight is minimal. Recent reports from other regions in Nigeria and across Africa indicate ongoing challenges related to contamination and AMR; however, data specific to northeastern Nigeria remain scarce [8,9]. This study aims to investigate the prevalence, geographic distribution, and AMR patterns of *Salmonella* spp., *Shigella* spp., and *L. monocytogenes* in poultry feeds and RTE foods across seven standardized markets in Maiduguri. By assessing contamination in poultry feeds (upstream) in conjunction with RTE foods (downstream), as well as associated grouped food categories and hygiene indicators (*coliforms*), this research intends to provide critical evidence that can inform targeted food safety interventions and One Health strategies in this underserved region.

## 2.0 Materials and Methods

### 2.1 Study Design and Setting

This cross-sectional study was conducted between 3 January, 2022 and 16 October, 2024 within the Maiduguri metropolis, situated in Borno State, northeastern Nigeria. Maiduguri serves as the capital city of Borno State, a region adversely affected by conflict, wherein a considerable portion of the population relies on informal poultry production and the sale of ready-to-eat (RTE) foods. Seven established markets were selected due to their substantial distribution of poultry feed and vending of RTE foods: Bama Road, Bulumkutu, Monday Market, Custom Market, Damboa Road, Moduganari, and Baga Road. The study design enabled a simultaneous evaluation of pathogen prevalence in poultry feeds (upstream) and RTE foods (downstream) in order to assess the associated risks of farm-to-fork transmission. This cross-sectional study is done in accordance with the STROBE rules/guidelines for observational studies

### 2.2 Sample Collection

A total of 120 poultry feed samples were collected from vendors and farms across the seven designated markets. The feeds were sampled aseptically from various brands and batches, ensuring approximately equal representation from each location (n=28 for Bama Road, n=18 for Bulumkutu, n=8 for Monday Market, n=19 for Custom Market, n=12 for Damboa Road, n=3 for Moduganari, and n=14 for Baga Road). For the analysis of *Salmonella* spp. and *Shigella* spp., approximately 251 RTE food samples were obtained, while 120 samples specifically for *Listeria monocytogenes* were procured from the same markets. The RTE food samples were categorized into specific groups: fruits (∼69), meats (n=28), milk and milk products (n=34), and vegetables (n=17). All samples were collected in sterile containers, transported on ice to the laboratory, and processed within four hours to ensure accuracy and reliability of the results.

### 2.3 Microbiological Analysis

Standard methodologies established by ISO and the FDA-BAM were employed for the processes of isolation and identification, with minor modifications implemented due to resource limitations within the study environment. For the isolation of *Salmonella* spp., 25 grams from each sample underwent initial pre-enrichment in buffered peptone water, followed by selective enrichment in Rappaport-Vassiliadis soy broth and tetrathionate broth. This was succeeded by plating on xylose lysine deoxycholate (XLD) and brilliant green agar. Suspected colonies were subsequently confirmed through biochemical methods. For *Shigella* spp., similar enrichment protocols were utilized (adapted from FDA BAM Chapter 6 and ISO 21567), with plating on MacConkey agar and *Salmonella*-*Shigella* agar, ensuring that biochemical confirmation followed. *Listeria monocytogenes* and other *Listeria spp*. were enriched in modified Listeria enrichment broth obtained from the University of Vermont, plated on Oxford and PALCAM agar, and confirmed via biochemical tests and motility assessments.

*Coliforms* were assessed through direct plating on MacConkey agar for colony counts, with presumptive *coliforms* identified as pink or red lactose-fermenting colonies; further characterization was undertaken using biochemical testing. Isolates underwent purification on Brain Heart Infusion and Nutrient agar.

### 2.4 Antimicrobial Susceptibility Testing

The assessment of antimicrobial resistance was conducted for isolates of *Salmonella* spp. (12 feeds, 3 ready-to-eat (RTE) samples) and *L. monocytogenes* (3 RTE samples) utilizing the Kirby-Bauer disk diffusion method on Mueller-Hinton agar, in accordance with Clinical and Laboratory Standards Institute (CLSI) guidelines. The antibiotics examined included tetracycline, fluoroquinolones (specifically ciprofloxacin, enrofloxacin, pefloxacin, ofloxacin, and levofloxacin), aminoglycosides (such as gentamicin, neomycin, and netilmicin), β-lactams (comprising amoxicillin and amoxicillin/clavulanic acid), in addition to several other agents (including ceftriaxone, trimethoprim-sulfamethoxazole, clindamycin, cefoxitin, erythromycin, doxycycline, and colistin). Zones of inhibition were classified as resistant, intermediate, or sensitive based on CLSI breakpoints.

### 2.5 Data Analysis

Prevalence was calculated as the percentage of positive samples; with 95% confidence intervals derived using the Wilson score method for binomial proportions. Descriptive analyses were performed to compare location-specific and food-category prevalence. Antimicrobial resistance (AMR) patterns were summarized as percentages of resistance corresponding to each antibiotic group. Due to the limited number of isolates available for AMR analysis, statistical significance was not determined. Data analysis was executed utilizing SPSS (version 23.0).

### 2.6 Ethical Considerations

The study protocol underwent a comprehensive review and received official approval from the Ministry of Health and Human Services (SHREC No. 021/2024, 0100/2021, and 138/2022) Health Research Ethics Committee. No human or animal subjects were directly involved in this research; all sampling was conducted on commercial feeds and available foods, with explicit consent obtained from vendors. Additionally, no personal data were collected during the course of this study.

## 4.0 RESULTS

### 4.1 Location-Specific Prevalence of Pathogens in Poultry Feeds and Ready-to-Eat (RTE) Foods

*Salmonella* spp. showed the highest overall prevalence in poultry feeds at 10.0% (95% CI: 5.8–16.8), with clear market hotspots. The highest contamination occurred in Monday Market (25.0%) and Bama Road (25.0%). In RTE foods, prevalence was lower at 2.3% (95% CI: 0.8–6.7) and limited to Bulumkutu fruits (pawpaw and watermelon) and Custom Market meat. *Shigella* spp. was detected in 3.33% (95% CI: 1.3–8.1) of poultry feeds, with the highest rates in Bulumkutu (11.1%) and Damboa Road (8.3%). Only one RTE isolate (0.76%) was recovered from pineapple (unspecified market), with an additional positive in Custom Market RTE. *Listeria monocytogenes* was absent from all feed samples (N/A) but present exclusively in RTE foods at 2.5% (95% CI: 0.8–7.1). The highest RTE rates were recorded in Baga Road (14.29%) (meat and sala sour milk) and Monday Market (12.50%).

**Table 4.1.**
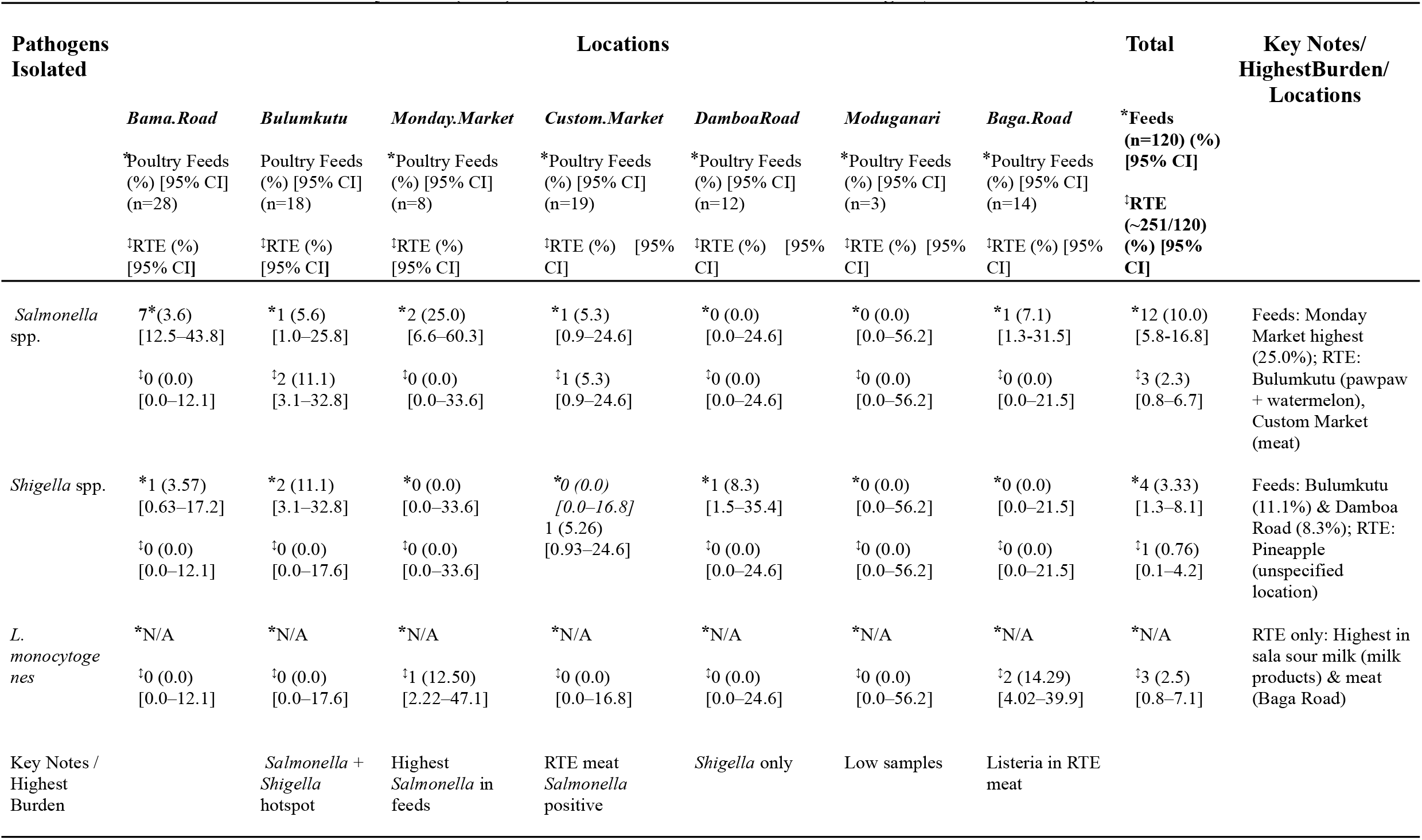
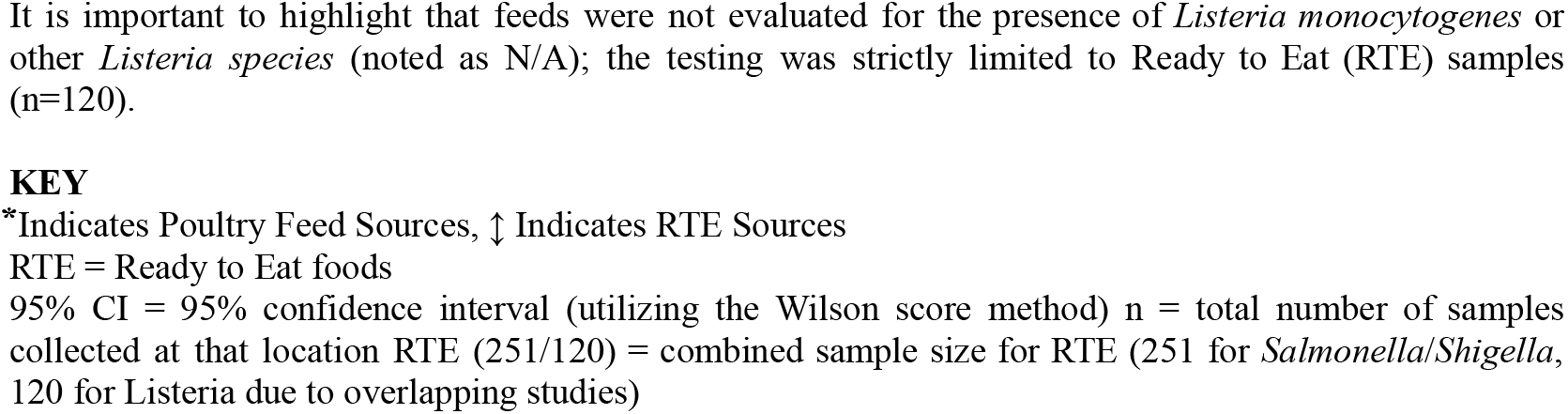
Location-specific prevalence of *Salmonella* spp., *Shigella* spp., and *Listeria monocytogenes* in poultry feeds and ready-to-eat (RTE) vended foods across markets in Maiduguri, northeastern Nigeria.

### 4.2 Prevalence by Grouped RTE Food Category

The prevalence of pathogens was further categorized according to the RTE food group (fruits ≈69 samples, meats n=28, milk/milk products n=34, vegetables n=17) to identify high-risk food categories and to conduct comparisons with poultry feeds (n=120). Contamination by *Salmonella* spp. in RTE foods was confined to fruits (2.9%, 95% CI 0.8–9.9; specifically pawpaw and watermelon) and meats (3.6%, 95% CI 0.6–17.2). The detection of *Shigella* spp. was limited to fruits (1.4%, 95% CI 0.2–7.8; found in pineapple). *Listeria monocytogenes* was tested exclusively in RTE samples and was detected at a prevalence of 3.6% in meats and 5.9% in milk products (the highest prevalence observed in sala sour milk), thereby confirming an elevated risk associated with animal-derived RTE categories in the markets of Maiduguri.

**Table 4.2.**
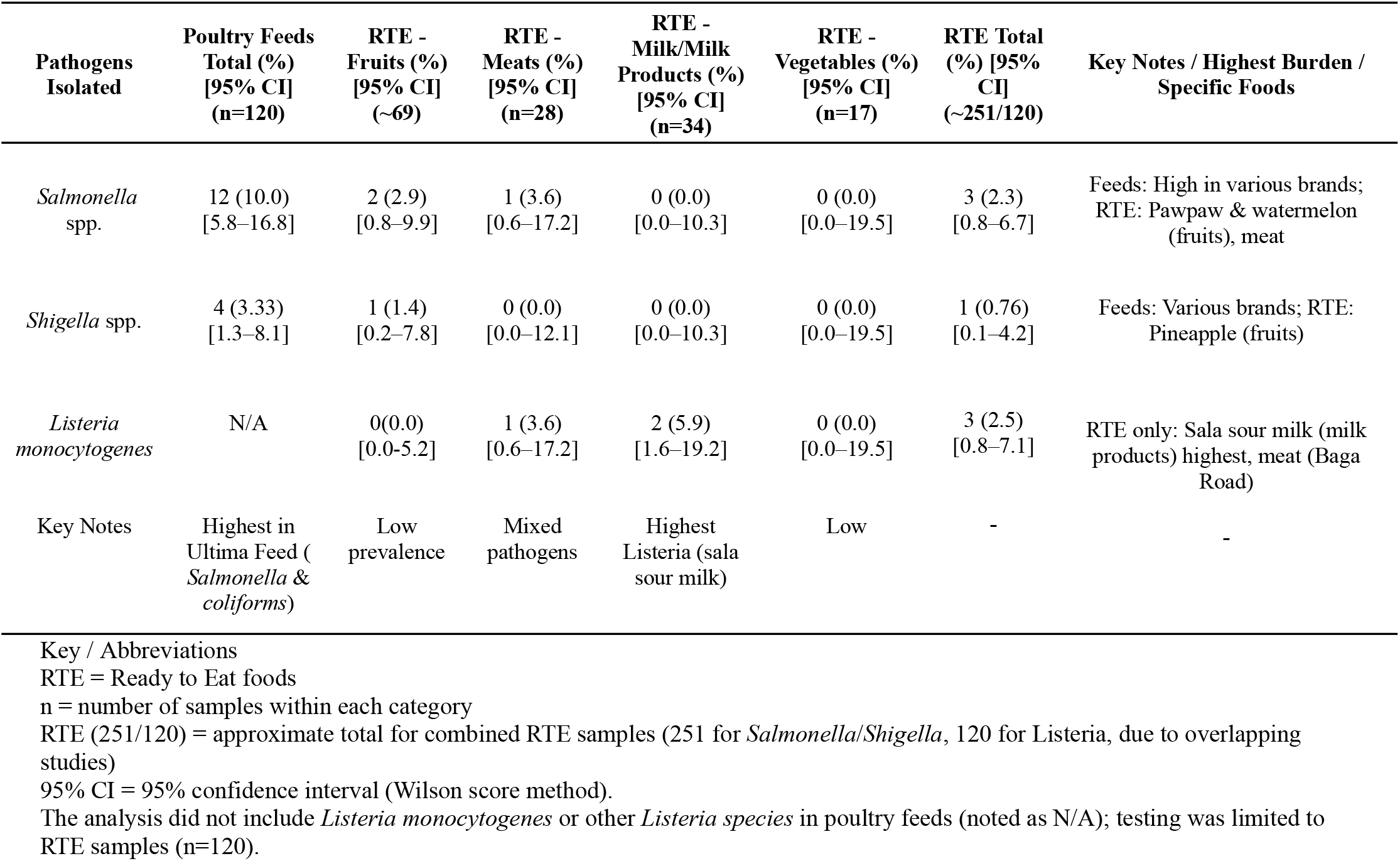
Prevalence of *Salmonella* spp., *Shigella* spp., and *Listeria monocytogenes* in poultry feeds, categorized by Ready-to-Eat (RTE) food types (fruits, meats, milk products, vegetables) in Maiduguri, northeastern Nigeria.

### 4.3 Prevalence of *Coliforms* and Non-Monocytogenes *Listeria species* by Location

*Coliforms* were quantified as a general indicator of hygiene, while non-monocytogenes *Listeria species* (L. welshimeri, L. ivanovii, L. innocua, L. grayi) were assessed exclusively in RTE samples (poultry feeds were excluded from testing). The prevalence of *coliforms* in poultry feeds was significantly high at 60.0% (95% CI 50.9–68.5), with the highest rate recorded in Bama Road at 75.0%. In contrast, the prevalence of *coliforms* in RTE foods decreased to 13.7% (95% CI 8.8–20.6). Non-monocytogenes *Listeria species* were detected solely in RTE samples, with a prevalence rate of 8.3% (95% CI 4.6–14.6). The highest occurrence was noted in Custom Market at 26.3%, followed by Bulumkutu (11.1%), Monday Market (12.5%), and Baga Road (14.3%). These findings indicate considerable upstream hygiene deficiencies and environmental contamination resulting from post-harvest practices at vending sites.

**Table 4.3.** Prevalence of *coliforms* and non-monocytogenes *Listeria species* in poultry feeds and RTE foods by standardized location in Maiduguri, northeastern Nigeria.

| Pathogens Isolated | Locations |  |  |  |  |  |  | Total |
| --- | --- | --- | --- | --- | --- | --- | --- | --- |
|  | *Bama Road* | *Bulumkutu* | *Monday Market* | *Custom Market* | *Damboa Road* | *Moduganari* | *Baga Road* | Feeds |
|  | Poultry Feeds (%) [95% CI] (n=28) | Poultry Feeds (%) [95% CI] (n=18) | Poultry Feeds (%) [95% CI] (n=8) | Poultry Feeds (%) [95% CI] (n=19) | Poultry Feeds (%) [95% CI] (n=12) | Poultry Feeds (%) [95% CI] (n=3) | Poultry Feeds (%) [95% CI] (n=14) | (n=120) (%) [95% CI] |
|  | RTE (%) [95% CI] | RTE (%) [95% CI] | RTE (%) [95% CI] | RTE (%) [95% CI] | RTE (%) [95% CI] | RTE (%) [95% CI] | RTE (%) [95% CI] | RTE (~251/120) (%) [95% CI] |
| *Coliforms species* | Feeds 21 (75.0) [57.1–87.3] | Feeds 11 (61.1) [38.6–79.7] | Feeds 5 (62.5) [30.6–86.3] | Feeds 6 (31.6) [15.4–54.0] | Feeds 7 (58.3) [32.1–80.7] | Feeds 2 (66.7) [20.8–93.9] | Feeds 10 (71.4) [45.4–88.3] | Feeds 72 (60.0) [50.9–68.5] |
|  | RTE: 3 (10.7) [3.7–27.2] | RTE: 2 (11.1) [3.1–32.8] | RTE: 1 (12.5) [2.2–47.1] | RTE: 2 (10.5) [2.9–31.4] | RTE: 1 (8.3) [1.5–35.4] | RTE: 0 (0.0) [0.0–56.2] | RTE: 2 (14.3) [4.0–39.9] | RTE: 18 (13.7) [8.8–20.6] |
| Other *Listeria spp.* (*L. welshimeri*, *L. ivanovii*, *L. innocua*, *L. grayi*) | Feeds N/A | Feeds N/A | Feeds N/A | Feeds N/A | Feeds N/A | Feeds N/A | Feeds N/A | Feeds N/A |
|  | RTE: 0 (0.0) [0.0–12.1] | RTE: 2 (11.1) [3.1–32.8] | RTE: 1 (12.5) [2.2–47.1] | RTE: 5 (26.3) [11.5–49.0] | RTE: 0 (0.0) [0.0–24.6] | RTE: 0 (0.0) [0.0–56.2] | RTE: 2 (14.3) [4.0–39.9] | RTE: 10 (8.3) [4.6–14.6] |
| Key Notes | High *coliforms* (75.0%) | *Salmonella* and *Shigella* hotspot | Highest *Salmonella* in feeds | RTE meat *Salmonella* positive | *Shi gella* only | Low samples | *Listeria* in RTE meat and sour milk | — |

Feeds were not assessed for the presence of *Listeria monocytogenes* or other *Listeria species* (designated as N/A); testing was confined to ready-to-eat (RTE) samples (n=120).
Key / Abbreviations
\*= Sources of Poultry Feeds ↑ = Sources of RTE 95% CI = 95% confidence interval (utilizing the Wilson score method) n = number of samples per location RTE (251/120) = estimated cumulative sample size for RTE (251 for *Salmonella* /*Shigella*, 120 for *Listeria* based on overlapping studies).

### 4.4 Antimicrobial Resistance Patterns

Antimicrobial resistance (AMR) was evaluated for *Salmonella* spp. (15 isolates), *Shigella* spp. (5 isolates), and *L. monocytogenes* (3 isolates from RTE only) through the disk diffusion method in accordance with CLSI guidelines. *Salmonella* exhibited universal resistance to tetracycline (100%) and displayed significant multi-drug resistance in feeds. *Shigella* demonstrated complete resistance to tetracycline while maintaining full sensitivity to fluoroquinolones. *L. monocytogenes* (limited to RTE) was entirely susceptible to tetracycline but exhibited 100% resistance to both fluoroquinolones and cephalosporins. These distinct antimicrobial resistance patterns provide compelling evidence of matrix-dependent AMR selection and the transmission of tetracycline resistance from farm to fork in this conflict-affected context.

**Table 4.4.** Antimicrobial resistance patterns of *Salmonella* spp. and *Listeria monocytogenes* isolate from poultry feeds and RTE foods in Maiduguri, northeastern Nigeria.

| Pathogens Isolated | No. of Isolates | Antibiotic Agent Groups |  |  |  | Other Notable Patterns | Findings/Implications |
| --- | --- | --- | --- | --- | --- | --- | --- |
|  |  | Tetracyclin | Fluoroquinilone | Aminoglycosides | Penicillin/B-lactams |  |  |
| *Salmonella* Species | 15 | 100% Resistant | 83.33% Resistant (CIP, ENR, PEF) | 0% Resistant (GEN, NEO) | 100% Resistant (AMX) | 66.67% Resistant (DOX); 50% Resistant (Colistin) | High multi-drug resistance (MDR) in feeds – alarming upstream reservoir; universal tetracycline resistance shared with RTE |
| *Shigella* species | 5 | 100% Resistant | 0% Resistant (CIP, OFX, LEV) | 66.7% Resistant (GEN, NTM) | Variable (66.7% AMC) | Partial resistance to ceftriaxone, SXT, pefloxacin | Tetracycline resistance preserved (farm-to-fork evidence); fluoroquinolone sensitivity retained in RTE (contrast to feeds) – potential dilution or selection downstream |
| *Listeria monocytogenes* | 3 | 100% Sensitive | 100% Resistant (CIP) | Not tested | Variable (66.7% AMC) | 100% Resistant (CD, COX); 66.7% Sensitive (E) | High fluoroquinolone and cephalosporin resistance – concerning for treatment; tetracycline sensitivity contrasts *Salmonella* pattern; RTE-specific risk in dairy/meat |

## 5.0 Discussion

This study represents one of the initial investigations focused on the multi-pathogen assessment of farm-to-fork transmission and antimicrobial resistance (AMR) relating to *Salmonella* spp., *Shigella* spp., and *Listeria monocytogenes* in poultry feeds and ready-to-eat (RTE) foods offered in conflict-affected Maiduguri, Borno State, northeastern Nigeria. The results indicate a distinct upstream-to-downstream gradient, with a prevalence of *Salmonella* spp. recorded at 10.0% (95% CI 5.8–16.8) in feeds, particularly in hotspots such as Monday Market and Bama Road, where the prevalence reached 25.0%. In contrast, the prevalence of *Salmonella* spp. in RTE foods was noted at 2.3% (0.8–6.7). *Shigella* spp. was detected in 3.33% (1.3–8.1) of feeds but was observed at only 0.76% in RTE foods. *L. monocytogenes* was exclusively found in RTE samples, occurring at a prevalence of 2.5% (0.8–7.1), primarily concentrated in sala sour milk and meat sourced from Baga Road and Monday Market.

The observed prevalence of 10.0% for *Salmonella* in poultry feeds is lower than the 41.2% reported in retail poultry meat from Benin City, Nigeria, yet it aligns with the pooled African estimate of 14.4% in informal poultry systems. The identified hotspots at Monday Market and Bama Road reflect market-level amplification that is indicative of the disrupted supply chains prevalent in northeastern Nigeria. This finding corresponds with observations from commercial farms in Oyo and Ibadan, where inadequate storage conditions and cross-contamination exacerbate contamination issues. The prevalence of 2.3% in RTE foods is significantly lower, suggesting potential dilution during the processing and vending phases; however, the presence of pathogens in pawpaw, watermelon, and meat aligns with transmission patterns reported in traditional markets in Ghana.

*Shigella* spp. exhibited a predominantly feed-based presence (3.33%), which contrasts sharply with the higher rates (5–10%) recorded in RTE foods across street vendors in Kenya and Ethiopia. The low detection rate in RTE foods (0.76%, specifically in pineapple) alongside preserved tetracycline resistance indicates limited downstream transmission in this context, corresponding with recent meta-analyses from Africa that emphasize the predominance of human handling over feed-related sources.

*L. monocytogenes* was absent in feeds (not tested) but was identified at a rate of 2.5% in RTE foods, with the highest frequencies observed in dairy (sala = sour milk) and meat. This occurrence is typical of post-harvest contamination found in refrigerated RTE products. The exclusive detection of *L. monocytogenes* in RTE and an additional 8.3% of non-monocytogenes *Listeria spp*. aligns with reviews underscoring significant risk points within informal vending environments for psychrotrophic pathogens.

The high levels of *coliforms* detected in feeds (60.0%, with a peak of 75.0% in Bama Road) highlight notable upstream hygiene deficiencies, comparable to the 50–80% ranges documented in Nigerian poultry feeds. The subsequent reduction to 13.7% in RTE signifies some improvement during vending, though it continues to indicate a considerable public health risk.

Antimicrobial resistance patterns reveal matrix-dependent selection alongside farm-to-fork transmission dynamics. The total of 15 *Salmonella* isolates demonstrated 100% resistance to tetracycline and 83.33% resistance to fluoroquinolones in feeds, whereas RTE isolates maintained tetracycline resistance yet exhibited a loss of fluoroquinolone resistance—reflecting similar trends observed within Iraqi and Nigerian poultry chains where the application of tetracycline in feeds sustains resistance. The five isolates of *Shigella* displayed uniform tetracycline resistance while remaining sensitive to fluoroquinolones, further supporting the notion of transmission of this resistance. In contrast, L. monocytogenes, found exclusively in RTE, showed 100% sensitivity to tetracycline but complete resistance to fluoroquinolones and cephalosporins, thereby highlighting notable pathogen-specific divergence in AMR and raising critical treatment considerations for listeriosis among vulnerable populations.

The findings presented are particularly concerning within the context of conflict-affected Maiduguri, where factors such as displacement, informal markets, and insufficient surveillance contribute to increased food insecurity and pathogen exposure among internally displaced persons (IDPs), children, and other vulnerable populations. The prevalence of malnutrition, in conjunction with disrupted health services, further heightens the risks associated with foodborne illnesses and resistant infections.

This study is subject to several limitations, including its cross-sectional design, which restricts the ability to draw causal inferences regarding transmission dynamics. Employing molecular typing methodologies, such as serovar identification and whole-genome sequencing for resistance genes, would enhance the evidentiary strength of the findings. Furthermore, antimicrobial resistance (AMR) testing was confined to 23 isolates, thereby limiting the potential for statistical comparisons and broader generalizability. Variations in sample sizes by pathogen and market, resulting from overlapping studies, additionally complicate the interpretation of results. Notably, vendor hygiene practices were not assessed directly, and the absence of testing for Listeria spp. in feeds may have resulted in overlooked upstream contamination sources.

In conclusion, this investigation highlights significant upstream contamination of Salmonella in poultry feeds, specific risks associated with ready-to-eat (RTE) products, the exclusive identification of *L. monocytogenes* in RTE items, and the transmission of tetracycline resistance within the farm-to-fork continuum in conflict-affected Borno State. The results present an urgent need for One Health interventions, which should encompass improved feed hygiene, the regulation of antimicrobial use within informal poultry systems, enhanced market surveillance, vendor training programs, and alignment with Nigeria’s National Action Plan on Antimicrobial Resistance (NAP-AMR 2.0, 2024–2028). This plan emphasizes multisectoral stewardship, agrifood system surveillance, and environmental considerations. Implementing these measures is essential for mitigating foodborne threats and addressing the emergence of AMR in underserved, conflict-impacted regions across sub-Saharan Africa.

## Declarations

## Acknowledgements

The authors sincerely thank the Department of Medical Microbiology, University of Maiduguri Teaching Hospital for technical support. We also appreciate the cooperation of feed vendors and RTE food sellers in Maiduguri markets.

## Authors’ Contributions

H.B.Ali: Conceptualization, supervision, data merging, formal analysis, writing – original draft, review and editing (corresponding author).

Jummai J.B: Investigation, methodology, data collection (poultry feeds study), laboratory analysis.

Barka A.M and Farida A.A: Investigation, methodology, data collection (RTE foods and Listeria study), laboratory analysis.

Isyaka M.Tom., Ajagbe J.M, Abubakar S.B, Usman M, and Yahaya U: Analysis, review and editing All authors read and approved the final manuscript.

### Funding

This research received no specific grant from any funding agency in the public, commercial, or not-for-profit sectors. It was self-funded by the corresponding author and students.

### Ethical Approval

Ethical clearance was obtained from the Borno State Ministry of Health and Human Services Medical Ethics Committee (SHREC No: 021/2024). All sampling was conducted with verbal consent from vendors. No human or animal subjects were directly involved. All sampling was done with verbal consent from the vendors. No human or animal subjects were directly involved in this study.

### Data Availability Statement

The datasets generated and analysed during the current study are The datasets generated and analyzed during this study are available within this article, and additional required information can be requested from the corresponding author.

## References

1. World Health Organization. WHO estimates of the global burden of foodborne diseases. Geneva: WHO; 2015.

2. Arias-Granda Z, Neuhofer ZT, Bauchet J, Ebner P, Ricker-Gilbert J. Foodborne diseases and food safety in sub-Saharan Africa: Current situation of three representative countries and policy recommendations. Afr J Agric Resour Econ. 2021;16(2):12–30.

3. World Food Programme. Northern Nigeria’s toxic mix of violence and hunger. Rome: WFP; 2025.

4. Jibril AH, Okeke IO, Dzikwi AA, et al. Prevalence and risk factors of Salmonella in commercial poultry farms in Nigeria. PLoS One. 2020;15(8):e0238190.

5. Ayele B, et al. Prevalence and Antimicrobial-Resistant Features of Shigella Species in East Africa from 2015–2022: A Systematic Review and Meta-Analysis. Infect Drug Resist. 2023;16:4567–4580.

6. Sibanda T, et al. Listeria monocytogenes at the food–human interface: A review of risk factors influencing transmission and consumer exposure in Africa. Int J Food Sci Technol. 2023;58(5):2345–2360.

7. Ramtahal MA, et al. A Public Health Insight into Salmonella in Poultry in Africa: A Review of the Past Decade: 2010–2020. Microb Drug Resist. 2022;28(3):345–360.

8. Cudjoe DC, et al. Food Safety in Sub-Sahara Africa, An insight into Ghana and Nigeria. Foods. 2022;11(24):4055.

9. Belina D, et al. Prevalence and epidemiological distribution of selected foodborne pathogens in human and different environmental samples in Ethiopia: a systematic review and meta-analysis. One Health Outlook. 2021;3:12.

10. Igbinosa EO, et al. Antimicrobial resistance and genetic characterisation of Salmonella enterica from retail poultry meats in Benin City, Nigeria. LWT. 2022;170:114057.

11. Kabeta T, et al. Prevalence and serotype of poultry salmonellosis in Africa: a systematic review and meta-analysis. Avian Pathol. 2024;53(4):301–315.

12. Orum TG, et al. Occurrence and antimicrobial susceptibility patterns of Salmonella species from poultry farms in Ibadan, Nigeria. Afr J Lab Med. 2022;11(1):1606.

13. Fadipe EO, et al. Prevalence of Salmonella outbreak in poultry farms: a comparative study of Osun and Ogun States, Nigeria. Int J Agril Res Innov Tech. 2025;15(1):115–126.

14. Hoffmann V, et al. Informal food systems and food safety in sub-Saharan Africa. Food Policy. 2019;85:1–10.

15. Moges M, et al. Sanitary condition and hygienic practice of street food vendors in selected towns of Ethiopia: A cross-sectional study addressing public health concern. J Agric Food Res. 2024;15:100912.

16. Onohuean H, et al. A scoping review of the prevalence of antimicrobial-resistant pathogens and signatures in ready-to-eat street foods in Africa. One Health. 2025;18:100456.

17. Dufailu OA, et al. Prevalence and characteristics of Listeria species from selected African countries. Foods. 2021;10(9):2123.

18. Centorotola G, et al. Listeria monocytogenes in ready to eat meat products from Zambia: phenotypical and genomic characterization of isolates. Foods. 2023;12(15):2890.

19. Adeyanju GT, Ishola O. Salmonella and Escherichia coli contamination of poultry meat from a processing plant and retail markets in Ibadan, Oyo State, Nigeria. Afr J Microbiol Res. 2014;8(4):398–405.

20. Jibril AH, et al. Association between antimicrobial usage and resistance in Salmonella from poultry farms in Nigeria. Antibiotics (Basel). 2021;10(8):975.

21. Igbinosa IH, et al. Identification and characterization of MDR virulent Salmonella spp isolated from smallholder poultry production environment in Edo and Delta States, Nigeria. PLoS One. 2023;18(2):e0281329.

22. World Health Organization. Global Antimicrobial Resistance and Use Surveillance System (GLASS). Geneva: WHO; 2025 [cited 2026 Mar 6]. Available from: https://www.who.int/initiatives/glass

